# Prevalence, Geographical Distribution and Determinants of Zero Dose Vaccination Status in Uttar Pradesh, India: An Analysis of The National Family Health Survey – 5 Data

**DOI:** 10.1101/2025.01.31.25321468

**Authors:** Prem Singh, Varun Sharma, Ashish Srivastava, Alan Noble John, Bhupendra Tripathi, Somesh Kumar

**Affiliations:** Jhpiego, New Delhi, India; Bill & Melinda Gates Foundation, New Delhi, India

**Keywords:** Determinants, Distribution, Healthcare Utilization, Immunization, Prevalence, Uttar-Pradesh, Zero-Dose Children

## Abstract

**Background:** Recent decades have witnessed considerable progress in immunization, leading to significant improvements in vaccination coverage worldwide. Despite these global achievements, regional disparities in vaccine uptake remain a challenge. In Uttar Pradesh (UP), India, there’s been a rise in the number of Zero-dose (ZD) children – those who have not received any routine vaccinations – over the past few years. This study delves into household-level data from the National Family Health Survey (NFHS-5, 2019-2021), aiming to uncover the prevalence and key factors contributing to the ZD status in UP.

**Methods:** Utilizing unit-level data from NFHS-5, we conducted a nested case-control analysis focusing on children aged 12-23 months. The primary outcome variable was ZD children, operationally defined as children who had not received the first dose of the pentavalent vaccine. The study focused on socio-demographic factors, maternal health, post-natal care, and systems’ engagement as potential predictors for ZD prevalence. These variables included type of residence, caste, religion, economic status, maternal education, place of delivery, antenatal care visits, and post-natal check-ups. Interactions with health workers and benefits received from community health programs were also considered.

**Results:** The prevalence of ZD children stands at 6.4% in UP. A significant urban-rural divide was observed, with 11.9% of urban children classified as ZD compared to 8.4% in rural areas. Bi-variate analysis revealed a significant association between ZD prevalence and residence type (χ2 =28.6421; p<0.01), with urban children being 37.1% more likely to be ZD than their rural counterparts at 24.5%. Economic status also showed a notable impact, with the poorest wealth quintile having 32.7% ZD children, significantly higher than the richer (23.4%) and richest (27.7%) quintiles. Maternal religion also showed significant association with ZD status, with 34.4% of children from Muslim households being ZD as compared to 24.9% in Hindu households. Maternal health indicators such as children of mothers with no antenatal care visits had 3.7 times higher odds (CI: 2.351 - 5.69; p<0.001) of being ZD compared to those whose mothers had 4+ visits. Engagement with health systems also influenced ZD rates; children from households that had not met community health workers were 1.6 times more likely to be ZD (OR: 1.566; CI: 1.135-2.16; p=0.006).

**Conclusions:** The higher prevalence of ZD children among specific groups underscores the need for targeted interventions. Strategies to address this issue must encompass an integration of maternal and child health services with existing immunization program.

## Introduction

Immunization is one of the most effective indicators to track that no one is left behind. Equity is embedded as a priority in global efforts to promote immunization. While tremendous progress has been made in the field of immunization in recent decades, with substantial improvements observed in vaccination coverage worldwide and a reduction in childhood morbidity and mortality due to vaccine-preventable diseases (VPD) [1], global inequities persist, reflected in the 25 million under-immunized and 18 million zero-dose children in 2021 [2]. “Zero-dose children” are those that have not received any routine vaccine. For operational purposes, Gavi defines zero-dose children as those who have not received the first dose of diphtheria-tetanus-pertussis containing vaccine (DTP1) [3].

The COVID-19 pandemic, associated disruptions, and COVID-19 vaccination efforts have strained health systems in 2020 and 2021, resulting in the number of zero-dose (ZD) children increasing by 5 million in 2021 compared with 2019 (13 million) and 25 million children missing out on vaccination, 6 million more than in 2019 and the highest number since 2008. [4] Over the years, much progress has been made in India to address immunization inequity and the proportion of ZD children in the country declined from 33.4% in 1992 to 10.1% in 2016 [5]. However, India continues to be the world’s largest birth cohort with substantial number of ZD children [6] As the DPT1 coverage declined from 94% in 2019 to 87% in 2020, the number of ZD children in the country doubled from 1.4 million in 2019 to over 3 million in 2020. As per the fifth round of the National Family Health Survey (NFHS-5, 2019-2021), 42 districts (out of 680 districts for which data is available in NFHS-5) had a prevalence of 15% and higher for ZD children, and eight of these 42 districts are in the state of Uttar Pradesh,[7] and the state is home to 53.0% of ZD children in the country, along with Bihar, and Maharashtra [8].

To improve immunization coverage and equity, the World Health Organization launched the Immunization Agenda 2030 (IA2030), with the goal of “leaving no one behind”, including a core objective to extend immunization services to reach ZD and under-immunized children and communities [9]. Aligned with IA2030, Gavi has launched its 5.0 Strategy with the goal of reducing the number of ZD children by 25% by 2025 and by 50% by 2030 [10]. In line with the above global commitments to reach ZD children, the Government of India has partnered with Gavi for focused support across 143 identified districts across 11 states.

These renewed efforts, both globally and in India, highlight the need to deep-dive into the existing data on ZD children to effectively reach them. Studies around the globe indicate that households with ZD children or Missed Communities are exposed to multiple deprivations, that is, the same households and communities are often exposed to several forms of scarcity, in terms of health services, nutrition, education, and the environment [11]. The accurate estimation of zero dose children remains unclear, with insufficient understanding of their identity, risk factors faced by them, their residential locations, and the persistent reasons behind the system’s failure to reach them. At the same time, there is a lack of micro-geographic level data about ZD children, along with their sociodemographic attributes and the factors that contribute to their risk.

This paper analyses the household-level data from NFHS-5 (2019-2021) for the state of Uttar Pradesh in India to determine the social and economic characteristics of households with ZD children, as well as the access of these households to other health services, with the aim of developing evidence-based recommendations to achieve high and equitable immunization coverage. The key research questions are: (a) What is the prevalence of zero-dose children across districts, and between urban and rural areas within those districts, in Uttar Pradesh? (b) How do the socio-demographic characteristics of households with zero dose child compare with households without a zero-dose child, in Uttar Pradesh? (c) How does the healthcare seeking behaviour during and immediately after pregnancy compare between mothers of zero dose children and mothers of non-zero dose children? And (d) Which factors are the determinants of zero dose vaccination status in Uttar Pradesh, India?

## Material and Methods

We used the unit-level data from the National Family and Health Survey – round 5 (NFHS-5). NFHS is a nationally representative survey of Indian households that provides essential information on health and family welfare. NFHS has been conducted periodically, with its first round (NFHS-I) conducted in 1992-93 and its latest round (NFHS-5) conducted in the year 2019-2021. All five rounds of NFHS were conducted under the guidance of the Ministry of Health and Family Welfare (MoHFW), Government of India, with the International Institute for Population Sciences (IIPS), Mumbai, designated as the nodal agency responsible for carrying out NFHS in all states and Union Territories (UTs) in India. The MoHFW, Government of India, provided the necessary funding for NFHS-5. NFHS-5 fieldwork in India was conducted in two phases: Phase-I from 17th June 2019 to 30th January 2020, which covered 17 states and 5 Union Territories (UTs), and Phase-II from 2nd January 2020 to 30th April 2021, which covered 11 states and 3 UTs. The survey was carried out by 17 Field Agencies, gathering information from a nationally representative sample, including 636,699 households, 724,115 women, and 101,839 men.

For the analysis in this paper, the unit level data from the NFHS-5 round is currently available through the DHS (Demographic and Health Surveys) and was sourced from the official DHS website (https://dhsprogram.com/data/available-datasets.cfm) after seeking proper approval.

We used the children below 5 years of age file (IAKR7DFL) and the Women unit-level data file (IAIR7DFL). These files were merged to obtain additional information, if available, on women respondents.

### Data and Sample Details

NFHS-5 provides health and family welfare data for 707 districts, 28 states, and 8 Union Territories (UTs) across India. To achieve this extensive coverage, the survey adopted a stratified two-stage sampling design. A sampling frame was established, referencing the census data from 2011, which served as the basis for the selection of Primary Sampling Units (PSUs). For rural areas, PSUs were represented by villages, while in urban areas, Census Enumeration Blocks (CEBs) served as the PSUs. Within each rural stratum, villages were chosen from the sampling frame using a probability proportional to size (PPS) approach. In urban areas, the Census Enumeration Block (CEB) information was obtained from the Office of the Registrar General and Census Commissioner, New Delhi. Before conducting the main survey, a comprehensive household mapping and listing exercise was carried out in every selected rural and urban Primary Sampling Unit (PSU). For PSUs with an estimated number of at least 300 households, they were further segmented into smaller segments, each comprising approximately 100-150 households. From each selected PSU or segment, two segments were randomly chosen for inclusion in the survey using systematic sampling with probability proportional to the segment size.

Consequently, an NFHS-5 cluster could be either a complete PSU or a segment of a PSU. In the second stage of sampling, within each selected rural and urban cluster, 22 households were randomly selected using systematic sampling. This multistage sampling approach ensured a well-structured and representative sample for the NFHS-5 survey. More detailed information on the sample design, questionnaire, pre-test, fieldwork, and data management are available elsewhere [12]. In the NFHS-5 survey, a total of 30,456 Primary Sampling Units (PSUs) were selected from 707 districts as of March 31st, 2017. Out of these, fieldwork was completed in 30,198 PSUs.

For the state of Uttar Pradesh, the NFHS-5 fieldwork was conducted in all 75 districts of the state. The survey took place in two phases: the first phase was conducted from 13th January 2020 to 21st March 2020, before the nation-wide lockdown during the pandemic, and the second phase was carried out from 28th November 2020 to 19th April 2021, after the lockdown. In Uttar Pradesh, a total of 73,976 households were selected for the survey, out of which 70,710 households were interviewed, resulting in a response rate of 97.3%. In total, 93,124 women aged 15-49 years were interviewed, with a response rate of 96.3%. Additionally, 12,043 men aged 15-54 years were interviewed, with a response rate of 88.6%.

### Variable of Interest

The focus of the paper is on the ZD children, which served as the outcome variable. To analyse this, we considered all children born to the participating women who were aged between 12 and 23 months at the time of the survey. Children who had passed away before the survey was conducted, were excluded from the analysis. In line with the IA2030 (Immunization Agenda 2030) definition, the term “zero-dose children” refers to those who do not have access to or are never reached by routine immunization services [13]. In this paper, we adopted the proposed IA2030 monitoring definition to define the variable of interest as a dichotomous (binary) variable. Accordingly, all surviving children aged between 12 and 23 months who did not receive the first dose of the pentavalent dose1 vaccine were categorized as “zero dose - ZD,” indicating that they had not received vaccination. On the other hand, children who had received the pentavalent dose1 vaccine were classified as “vaccinated”. During the NFHS surveys, a standardized procedure was employed to collect vaccination data, which was carried out by trained field personnel. The child’s primary caregiver, typically the mother, was interviewed, and requested to provide the child’s vaccination record. If the vaccination card was available, the vaccination data were recorded from the card directly. However, in cases where the vaccination card was not accessible or not present at the time of the interview, the vaccination data were gathered through caregiver.

### Explanatory Variables

We considered a range of socio-demographic, maternal health, post-natal care and systems’ engagement variables as potential predictors for zero-dose prevalence. The socio-demographic variables included were:

i. Type of residence: Categorized as urban or rural.
ii. Caste of mother: categorized into Scheduled Caste (SC), Scheduled Tribe (ST), Other Backward Class (OBC), and others (none of the mentioned castes).
iii. Religion of mother: Classified as Hindu or Muslim.
iv. Economic status (Wealth index quintiles): Categorized into five groups - poorest, poorer, middle, richer, and richest.
v. Highest education level of mother: Categorized as – no education, primary, secondary and higher

For the maternal health, we used two indicators – (a) place of delivery and (b) antenatal care visits during pregnancy. Place of delivery was classified into two categories viz. home and institutional delivery. Institutional delivery includes both public, private and voluntary sector facilities. Antenatal care visits during pregnancy were categorized into three categorize – (a) no visit; (b) 1-3 visits and (c) 4+ visits. For the post-natal care, we used post-natal check-up of baby within 2 months as an indicator. In addition to these variables, specific questions from the women’s questionnaire were used to determine the Systems’ engagement. The questions asked were as follows:

a. “In the last three months, have you met with an ANM (Auxiliary Nurse Midwife) or LHV (Lady Health Visitor)?”
b. “In the last three months, have you met with an *anganwadi* worker, ASHA (Accredited Social Health Activist), or other community health worker?”
c. “When you were pregnant with (child’s name), did you receive any benefits from the *anganwadi*/ICDS (Integrated Child Development Services) center?”

The responses to these questions were binary, with possible answers being “yes” or “no”.

### Analytical Approach

We used a nested-case-control method as the analytical approach for the analysis. The dataset included 6,553 unweighted observations and 6,412 weighted observations for the age group 12-23 months in Uttar Pradesh. To conduct the case-control analysis, we matched observations based on three variables: district, age of the child, and sex of the child. The aim was to identify case and control observations for the analysis.

Case observations were defined as children aged 12-23 months in Uttar Pradesh who had not received pentavalent dose 1 (ZD children). Contrary, control observations were defined as children aged 12-23 months in Uttar Pradesh who had received pentavalent dose 1 (vaccinated children). After matching the observations by district, age, and sex, we obtained a reduced dataset with 2,282 unweighted observations of children. Among these, there were 588 case observations (ZD children) and 1,694 control observations (vaccinated children) i.e. 1:3 ratio for case and control observations. This matching process allowed us to create a balanced dataset for further analysis, comparing the characteristics and factors associated with ZD prevalence in Uttar Pradesh.

We conducted a bivariate analysis to examine the association between the ZD prevalence and various other variables, including background characteristics, maternal health indicators, post-natal care and systems’ engagement. To measure the association between the ZD status and these variables, we used the chi-square test, that allowed us to determine if there is a significant association between ZD status and other categorical variables. The logistic regression analysis on the matched observations was conducted to explore the determinants of ZD prevalence among children aged 12-23 months in Uttar Pradesh. The outcome variable in this analysis is a dichotomous variable with values 0 and 1, representing the probability of a child either not receiving (1) or receiving (0) the pentavalent dose1 vaccine. The explanatory variables, on the other hand, are categorized into three main categories: Socio-demographic Characteristics, maternal health indicators including post-natal care and Systems’ Engagement. We used STATA 14.0 to conduct all statistical analyses.

### Ethical Statement

This paper used secondary data available in the public domain with all identifiable information removed; therefore, ethics committee approval was not obtained. The respondents in the NFHS undergo an informed consent process for participation in the survey after approval of the protocol by the institutional review board of the IIPS. Additionally, the protocol for the NFHS-5 survey, including the content of all the survey questionnaires, was approved by the IIPS Institutional Review Board and the ICF Institutional Review Board. The protocol was also reviewed by the U.S. Centers for Disease Control and Prevention (CDC).

## Results

Figure 1 shows the distribution of ZD prevalence across districts in Uttar Pradesh. Out of the 75 districts in Uttar Pradesh, about 32 districts have a proportion of ZD children that exceeds the state’s average of 9.1%, and around 53 districts surpass the national average of 6.4%. Kanpur Dehat district has the highest proportion of ZD prevalence (19.58%) and Agra district has the lowest proportion (0.81%).

**Figure 1:**
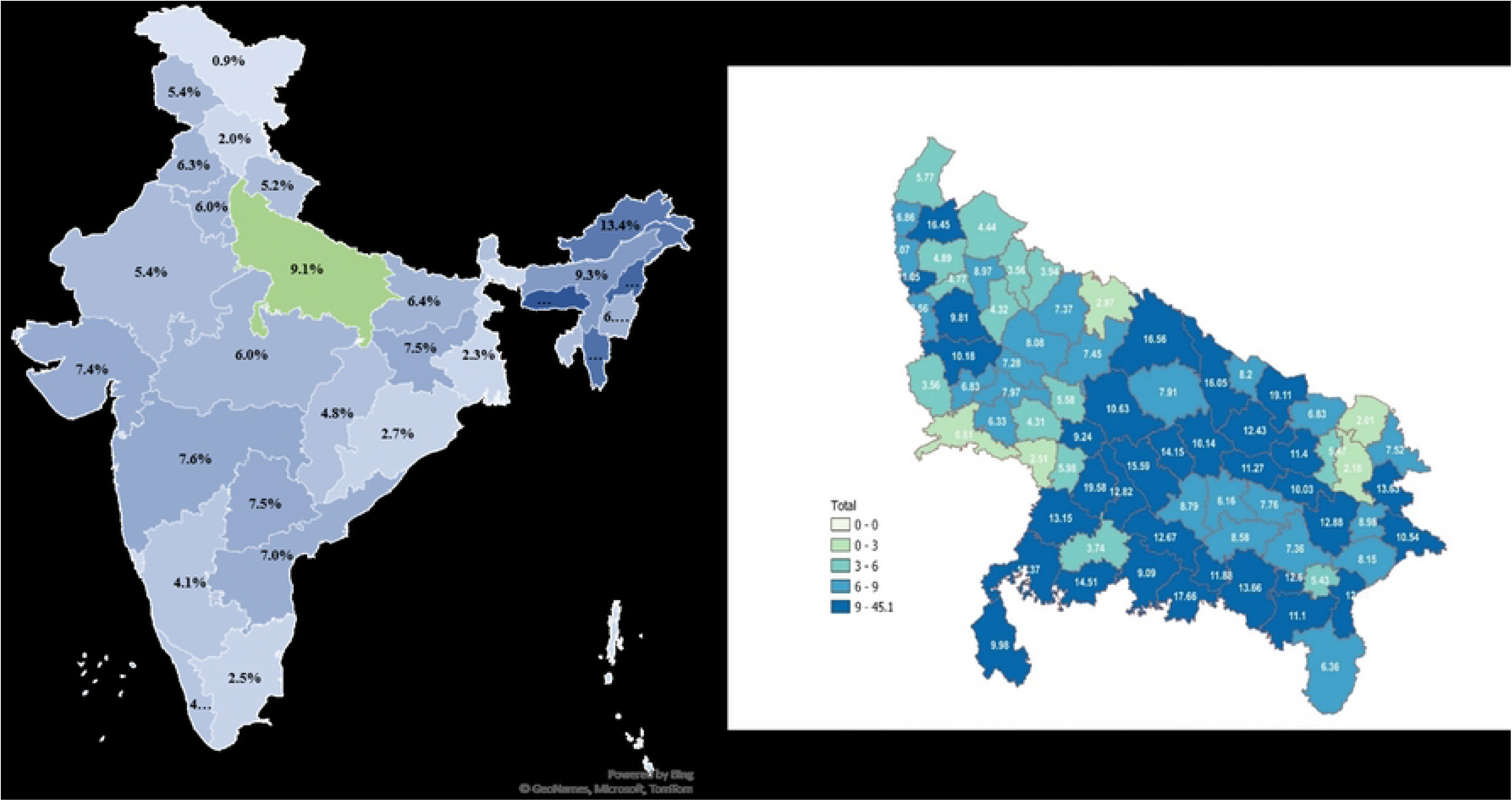
ZD Prevalence Among Children Aged 12-23 Months in India and Uttar Pradesh (NFHS-5)

Figures 2-A and 2-B present the prevalence of zero-dose children in urban and rural areas of Uttar Pradesh districts, respectively. In urban Uttar Pradesh, 11.9% of children are categorized as zero-dose, compared to 8.4% in rural Uttar Pradesh. The overall state average for zero-dose children is higher than just the rural average, suggesting that a significant number of zero-dose children reside in urban areas. Urban areas of districts like Balia (45.1%), Balrampur (44.2%), and Unnao (41.3%) have the highest proportions of zero-dose children, while in rural areas, the districts of Kanpur Dehat (21.2%), Chitrakoot (19.6%), and Muzaffarnagar (18.7%) lead in this category.

**Figure 2-A:**
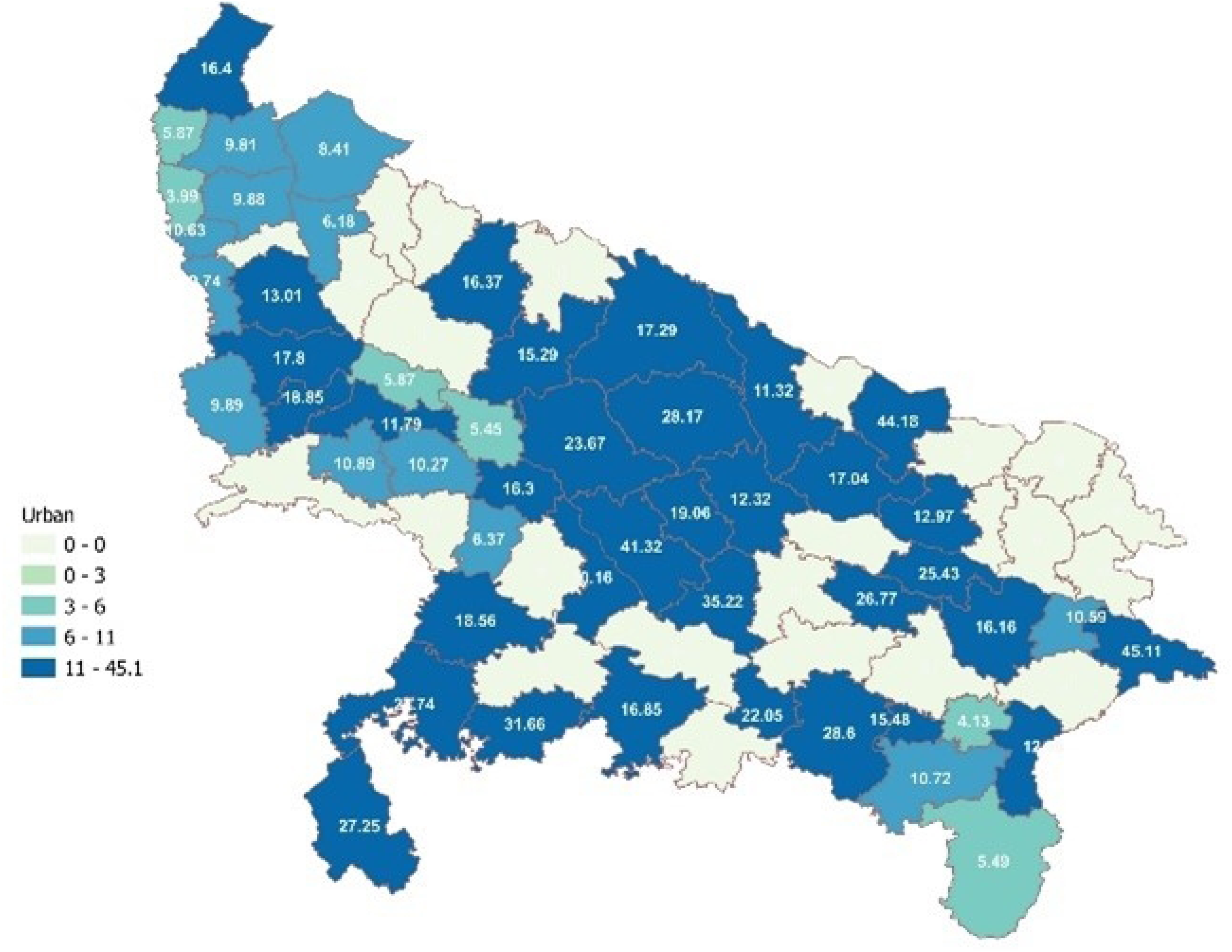
District Wise ZD Prevalence Among Children Aged 12-23 Months in Urban Uttar Pradesh (NFHS-5)

**Figure 2-B:**
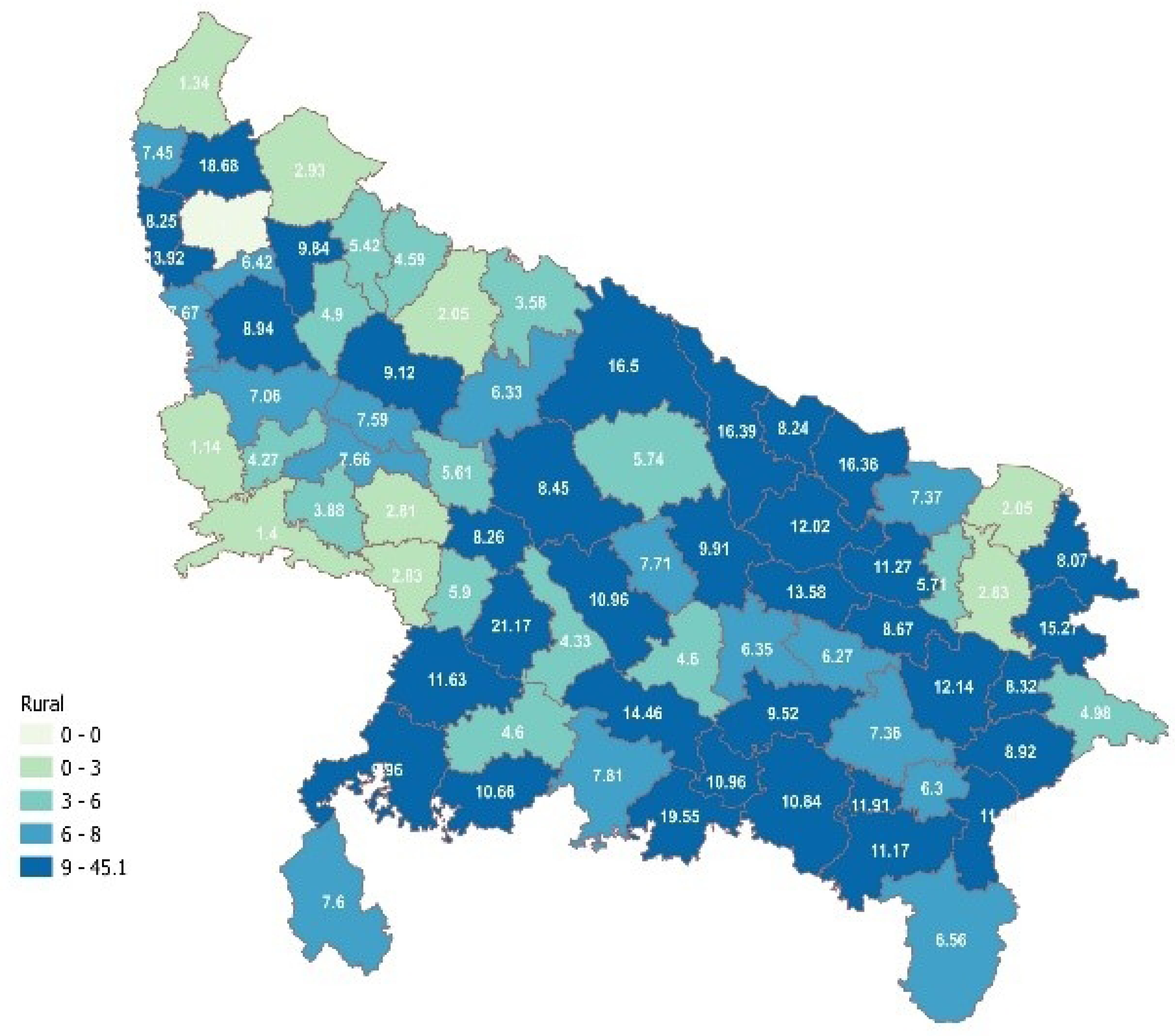
District Wise ZD Prevalence Among Children Aged 12-23 Months in Rural Uttar Pradesh (NFHS-5)

From the overall sample of children aged 12-23 months in Uttar Pradesh, we used a case-control design after matching observations as outlined in the methods section. The subsequent analysis in the paper is based on this analytical framework.

**Table 1** presents the association between background characteristics and ZD prevalence in Uttar Pradesh on matched cases. Bi-variate analysis suggests a strong association between type of residence and ZD prevalence (χ2 =28.6421; p<0.01). A large percentage of children aged 12-23 months in urban Uttar Pradesh were ZD children compared to children residing in rural areas (37.1% and 24.5% respectively). Caste-wise, no significant association was observed between caste of the mother and ZD prevalence, though compared to children belonging to Scheduled Caste (SC) and Other Backward Class (OBC), a large percentage of children from Scheduled Tribe (ST) were ZD children (35.4%). Religion was strongly associated with ZD prevalence (χ2 =17.3809; p=0.001). Among Muslims, 34.4 percent children aged 12-23 months were ZD children compared to 24.9 percent Hindu children. There appeared to be a strong association between the highest education level of the mother and ZD prevalence (χ2 = 19.611; p=0.002). One in every third child of mothers with no education (32.8%) was ZD child. For mother with higher education, it was 21.3 percent. A large percentage of children belonging to the poorest wealth quintile (32.7%) were ZD children compared to the children belonging to the richer (23.4%) and richest (27.7%) wealth quintile. The association between the economic status and ZD prevalence was statistically significant (χ2 = 16.188; p=0.027).

**Table 1:**
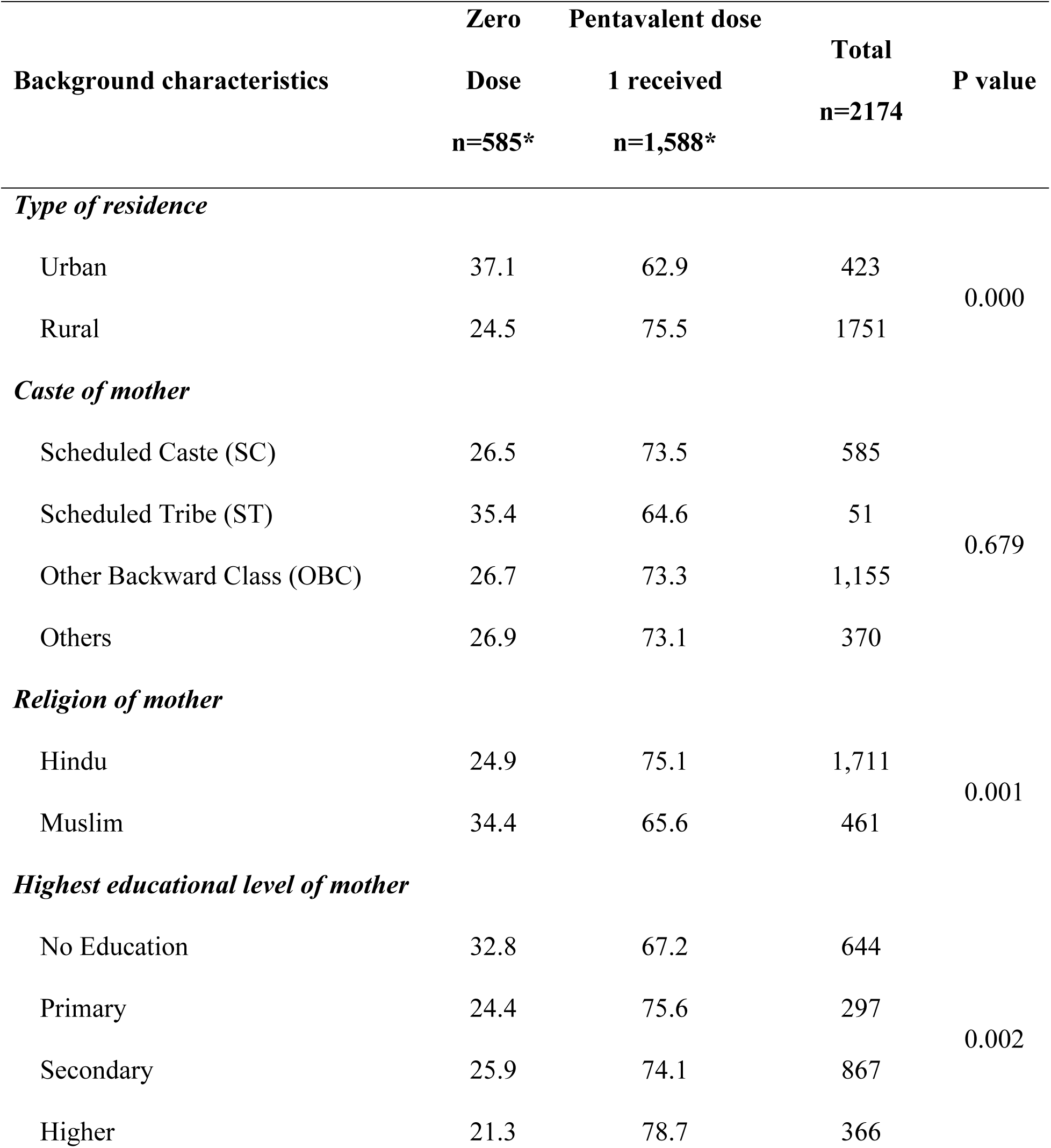
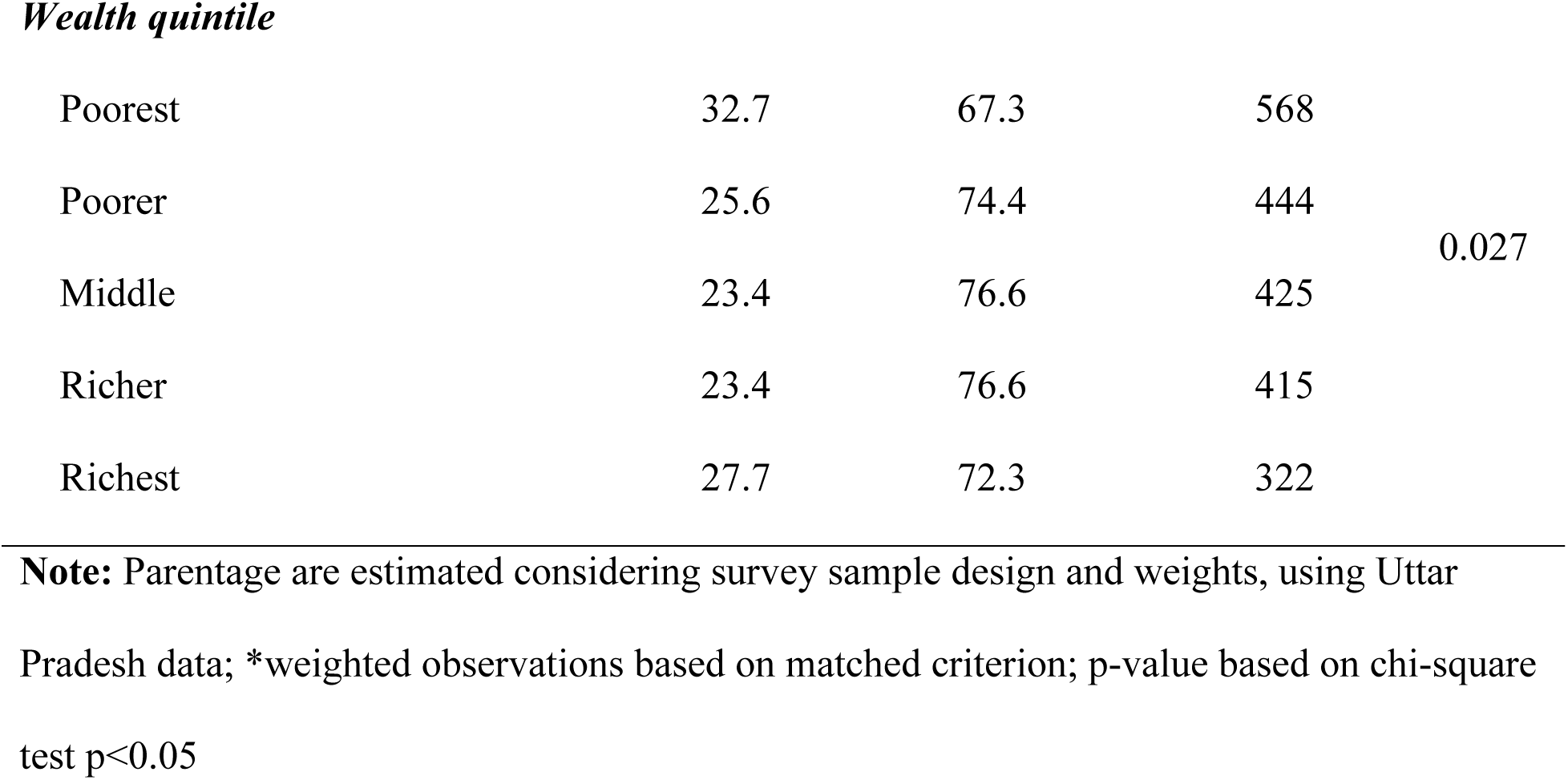
Sociodemographic characteristics and zero dose prevalence considering case-control sample in Uttar Pradesh.

| Background characteristics | Zero<br>Dose<br>n=585* | Pentavalent dose<br>1 received<br>n=1,588* | Total<br>n=2174 | P value |
| --- | --- | --- | --- | --- |
| *Type of residence* |  |  |  |  |
| Urban | 37.1 | 62.9 | 423 | 0.000 |
| Rural | 24.5 | 75.5 | 1751 |  |
| *Caste of mother* |  |  |  |  |
| Scheduled Caste (SC) | 26.5 | 73.5 | 585 | 0.679 |
| Scheduled Tribe (ST) | 35.4 | 64.6 | 51 |  |
| Other Backward Class (OBC) | 26.7 | 73.3 | 1,155 |  |
| Others | 26.9 | 73.1 | 370 |  |
| *Religion of mother* |  |  |  |  |
| Hindu | 24.9 | 75.1 | 1,711 | 0.001 |
| Muslim | 34.4 | 65.6 | 461 |  |
| *Highest educational level of mother* |  |  |  |  |
| No Education | 32.8 | 67.2 | 644 | 0.002 |
| Primary | 24.4 | 75.6 | 297 |  |
| Secondary | 25.9 | 74.1 | 867 |  |
| Higher | 21.3 | 78.7 | 366 |  |
| ***Wealth quintile*** |  |  |  |  |
| Poorest | 32.7 | 67.3 | 568 | 0.027 |
| Poorer | 25.6 | 74.4 | 444 |  |
| Middle | 23.4 | 76.6 | 425 |  |
| Richer | 23.4 | 76.6 | 415 |  |
| Richest | 27.7 | 72.3 | 322 |  |

**Note:** Parentage are estimated considering survey sample design and weights, using Uttar Pradesh data; \*weighted observations based on matched criterion; p-value based on chi-square test $p < 0.05$

### Association Between Zero-Dose Status and Maternal Health Indicators

We found a strong statistically significant association between maternal health indicators and pentavalent dose 1 uptake (Table 2). Children whose mothers had attended four or more ANC visits, a majority of them were vaccinated children (79.1%), compared to the children whose mothers did not attend any ANC visit (42.1%). A statistically significant association was observed between ANC visits and ZD prevalence (χ2 = 85.476, p<0.001). Children born at home were more likely to be ZD children compared to those who were born in institutional settings (37.4% vs. 24.9%, χ2 = 24.214, p<0.001). Similarly, a higher percentage of children who received a postnatal check-up within two months were also vaccinated (78.2%) compared to those who did not (68.9%) and the association was statistically significant (χ2 = 23.288, p<0.001).

**Table 2:** Association Between Pre and post Antenatal Care Indicators and Zero Dose Prevalence in Uttar Pradesh Considering Case-Control Sample.

| **Maternal Health and Post Natal**<br>**Care Indicators** | **Zero**<br>**Dose**<br>**n= 585** | **Pentavalent dose 1**<br>**received**<br>**n= 1588** | **Total**<br>**n=2174** | **P value** |
| --- | --- | --- | --- | --- |
| ***Number of ANC visits*** |  |  |  |  |
| No ANC visits | 57.9 | 42.1 | 127 | 0.000 |
| 1 to 3 ANC visits | 28.7 | 71.3 | 1,078 |  |
| More than equal to 4 visits | 20.9 | 79.1 | 969 |  |
| ***Place of delivery*** |  |  |  |  |
| Home | 37.4 | 62.6 | 347 | 0.000 |
| Institutional | 24.9 | 75.1 | 1,826 |  |
| ***Postnatal check of baby within two***<br>***months*** |  |  |  |  |
| No | 31.1 | 68.9 | 1,074 | 0.000 |
| Yes | 21.8 | 78.2 | 937 |  |

**Note:** Parentage are estimated considering survey sample design and weights, using Uttar Pradesh data; p-value based on chi-square test $p < 0.05$

### Association Between Zero-Dose Status and Systems’ Engagement

Around 30 percent of children whose mothers did not meet with an Auxiliary Nursing Midwifery (ANM) or Lady Health Visitor (LHV) in the last three months did not receive pentavalent dose 1, compared to 22 percent of children of mothers who met with an ANM or LHV in the last three months. The association between meeting with an ANM or LHV and pentavalent dose 1 uptake was statistically significant (χ2 =20.6174; p<0.001).

Likewise, among children whose mothers did not meet with an Anganwadi Worker (AWW), an Accredited Social Health Activist (ASHA), or other community health workers in the last three months, 35 percent were ZD children, compared to 21.5 percent for those who met any of the mentioned community health workers. The association between meeting with community health workers and pentavalent dose 1 uptake was statistically significant (χ2 =50.0327; p<0.001). The association between receiving benefits from the anganwadi while being pregnant and pentavalent dose 1 was statistically significant (χ2 = 67.941; p<0.001), given that 37.7 percent of children whose mothers did not receive any benefit from the anganwadi while being pregnant were ZD children compared to 21.5 percent of those who did receive any benefit from the anganwadi during pregnancy.

The analysis found significant associations between systems’ engagement indicators and the administration of pentavalent dose 1 (Table 3). Mothers who met with ANMs or LHVs, community health workers, or received benefits from the Anganwadi during pregnancy showed higher rates of pentavalent dose 1 uptake for their children.

**Table 3:**
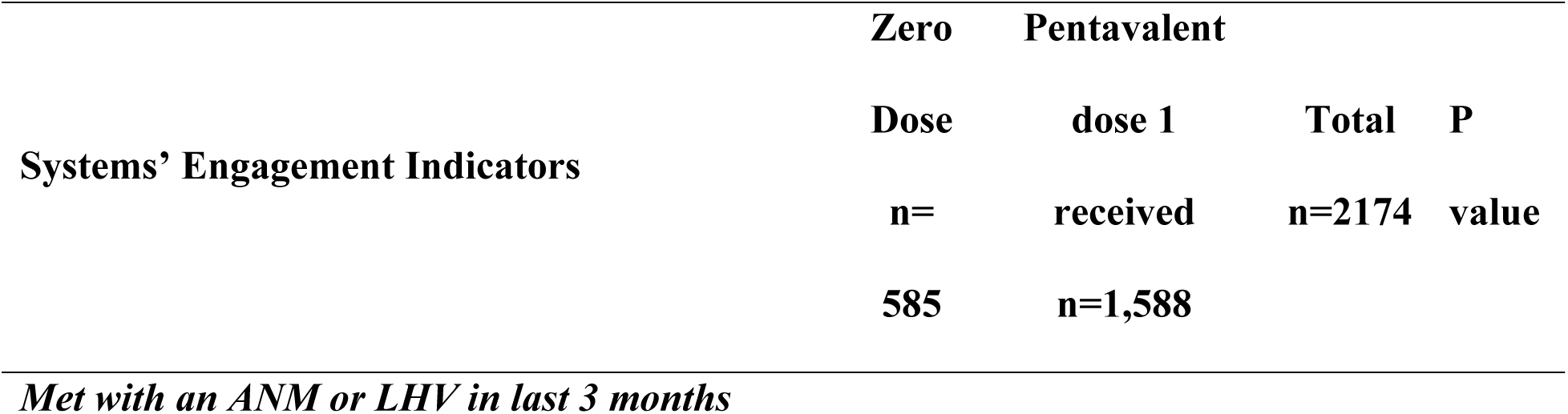
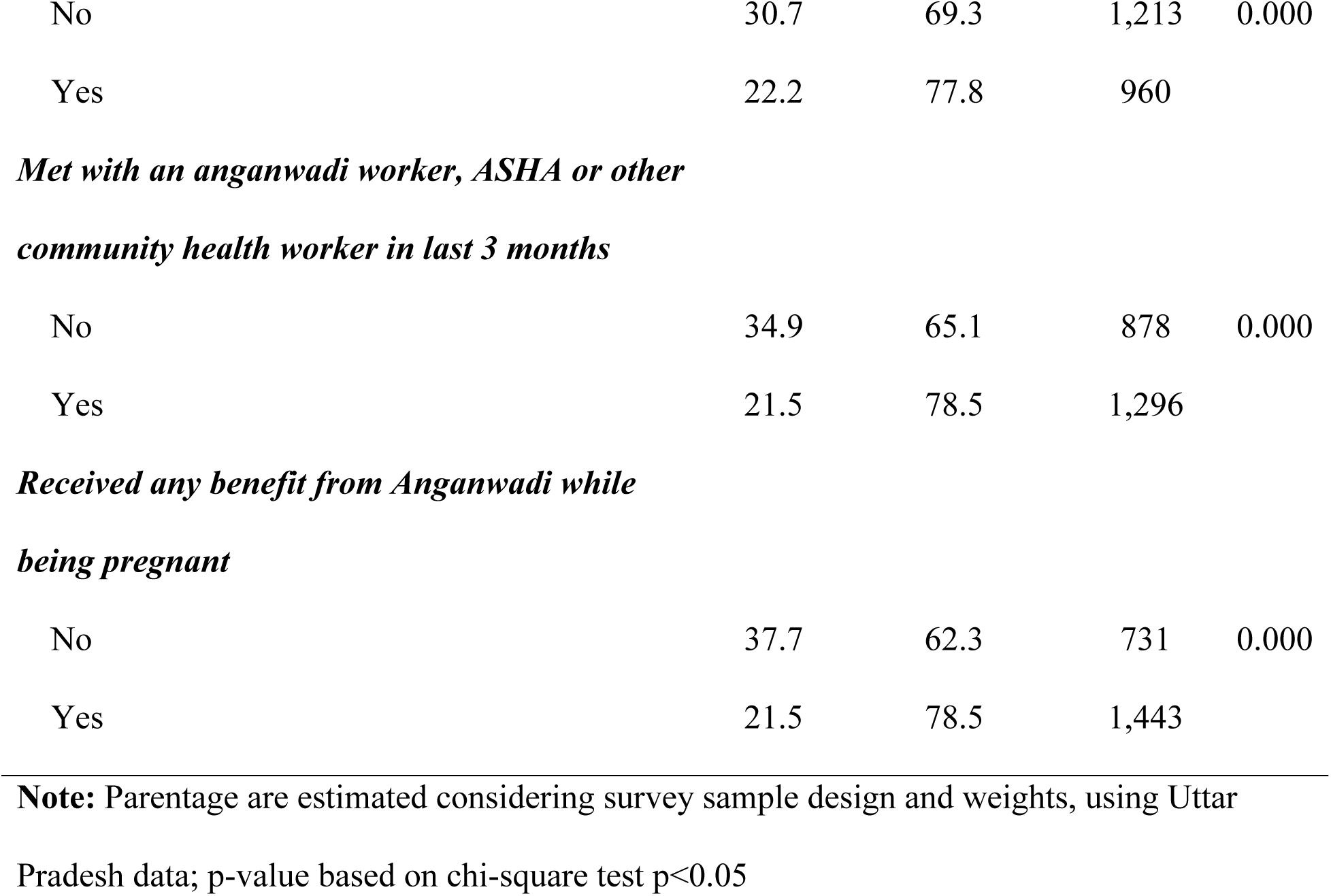
Systems’ engagement and zero dose prevalence in Uttar Pradesh considering case-control sample.

### Determinants of Zero-Dose Prevalence

Table 4 presents the results of logistic regression analysis describing the predictors of ZD prevalence among children aged 12-23 months in Uttar Pradesh. In the analysis we have reported both unadjusted and adjusted odds ratios for the variables. In this analysis, we have used ‘receiving pentavalent dose 1” as the reference category for the key interest variable ‘zero dose prevalence’. The analysis highlights the influence of socio-economic characteristics, households’ engagement with community outreach programs, and specific maternal health indicators as predictors of ZD prevalence. For predictors like caste and the mother’s educational status, the adjusted odds ratios do not show statistically significant results. For the rest predictor variables, such as place of residence, economic status, religion, household’ engagement with community outreach programs and pre/post-natal care indicators, a statistically significant association was found after adjusting for all confounders. Regarding the place of residence, after adjusting for all confounders, the odds of children in urban areas being zero dose were twice as compared to children in rural areas. Urban children are more likely to have not received pentavalent dose 1 compared to children residing in rural Uttar Pradesh. The odds of children residing in urban areas not receiving pentavalent dose 1 were 1.83 times higher i.e. 83% more likely (CI: 1.224 - 2.728) compared to children residing in rural areas. Caste-wise no significant association was observed. In terms of the economic status of households, there was a declining trend found as wealth increased from the poorest to richer classes. Children from poorer (OR: 0.683) and middle wealth quintiles (OR: 0.653) were around 30% less likely to fall into the zero-dose category than children from the poorest wealth quintile. Similarly, children in the richer wealth quintile (OR: 0.553) were 45% less likely to be in the zero-dose category than children from the poorest wealth quintile. However, for children in the richest wealth quintile, the trend differed; they were 40% less likely to be in the zero-dose category, but this association was not statistically significant (p value=0.09). While considering religion, after adjusting for all confounders, the odds of a child born in a Muslim household to be in the zero-dose category was 1.5 times (CI: 1.057 - 2.066; p=0.022) higher than the odds of child born in a Hindu household.

**Table 4:** Determinants of Zero-dose prevalence in Uttar Pradesh, India.

| Predictors of zero – dose<br>prevalence | Unadjusted |  | p-value | Adjusted |  | p-value |
| --- | --- | --- | --- | --- | --- | --- |
|  | OR | (95% CI) |  | OR | (95% CI) |  |
| Socio-demographic characteristics |  |  |  |  |  |  |
| Caste of mother |  |  |  |  |  |  |
| Scheduled Caste (SC) | 1.000 |  |  | 1.000 |  |  |
| Scheduled Tribe (ST) | 1.522 | [0.788 - 2.93] | 0.210 | 1.262 | [0.563 - 2.827] | 0.571 |
| Other Backward Class (OBC) | 1.011 | [0.772 - 1.323] | 0.936 | 1.031 | [0.774 - 1.372] | 0.834 |
| Other | 1.022 | [0.725 - 1.442] | 0.897 | 0.978 | [0.665 - 1.439] | 0.914 |
| Place of resident |  |  |  |  |  |  |
| Rural | 1.000 |  |  | 1.000 |  |  |
| Urban | 1.814 | [1.355 - 2.429] | 0.000 | 1.828 | [1.224 - 2.728] | 0.003 |
| *Educational status of mother* |  |  |  |  |  |  |
| No education | 1.000 |  |  | 1.000 |  |  |
| Primary | 0.660 | [0.464 - 0.939] | 0.021 | 0.8313 | [0.557 - 1.238] | 0.364 |
| Secondary | 0.715 | [0.552 - 0.925] | 0.011 | 1.005 | [0.748 - 1.35] | 0.973 |
| Higher | 0.553 | [0.399 - 0.766] | 0.000 | 0.7678 | [0.511 - 1.153] | 0.203 |
| *Household wealth index* |  |  |  |  |  |  |
| Poorest | 1.000 |  |  | 1.000 |  |  |
| Poorer | 0.708 | [0.515 - 0.972] | 0.033 | 0.683 | [0.492 - 0.947] | 0.023 |
| Middle | 0.628 | [0.458 - 0.86] | 0.004 | 0.653 | [0.458 - 0.93] | 0.018 |
| Richer | 0.629 | [0.443 - 0.894] | 0.010 | 0.553 | [0.37 - 0.826] | 0.004 |
| Richest | 0.789 | [0.549 - 1.134] | 0.201 | 0.648 | [0.392 - 1.072] | 0.092 |
| *Religion* |  |  |  |  |  |  |
| Hindu | 1.000 |  |  | 1.000 |  |  |
| Muslim | 1.578 | [1.204 - 2.07] | 0.001 | 1.478 | [1.057 - 2.066] | 0.022 |
| **Maternal health indicators** |  |  |  |  |  |  |
| *Antenatal care visits during pregnancy* |  |  |  |  |  |  |
| 4+visits | 1.000 |  |  | 1.000 |  |  |
| less than 4 visits | 1.526 | [3.393 - 7.987] | 0.000 | 1.587 | [1.23 - 2.047] | 0.000 |
| No visit | 5.206 | [1.209 - 1.927] | 0.000 | 3.658 | [2.351 - 5.69] | 0.000 |
| *Place of delivery* |  |  |  |  |  |  |
| Institutional | 1.000 |  |  | 1.000 |  |  |
| Home | 1.798 | [1.353 - 2.39] | 0.000 | 1.306 | [0.95 - 1.797] | 0.100 |
| **System's engagement** |  |  |  |  |  |  |
| *Met with an ANM or LHV in the last three months* |  |  |  |  |  |  |
| Yes | 1.000 |  |  | 1.000 |  |  |
| No | 1.551 | [1.24 - 1.942] | 0.000 | 0.9126 | [0.671 - 1.241] | 0.560 |
| *Met with an anganwadi worker*<br>*(AWW), ASHA or other community*<br>*worker in the last three months* |  |  |  |  |  |  |
| Yes | 1.000 |  |  | 1.000 |  |  |
| No | 1.955 | [1.56 - 2.45] | 0.000 | 1.566 | [1.135 - 2.16] | 0.006 |
| *Received any benefit from*<br>*Anganwadi while being pregnant* |  |  |  |  |  |  |
| Yes | 1.000 |  |  | 1.000 |  |  |
| No | 2.210 | [1.762 - 2.77] | 0.000 | 1.529 | [1.178 - 1.986] | 0.001 |
| **Postnatal indicator** |  |  |  |  |  |  |
| *Baby postnatal check within 2*<br>*months* |  |  |  |  |  |  |
| Yes | 1.000 |  |  | 1.000 |  |  |
| No | 1.621 | [1.283 - 2.048] | 0.000 | 1.243 | [0.97 - 1.593] | 0.085 |

**Note:** OR: Odds Ratio; p<0.05

For maternal health indicators, children whose mothers had no antenatal care (ANC) visits during pregnancy were more likely to be ZD children. Specifically, the odds that a child whose mother did not receive at least one ANC visit was 3.7 times (CI: 2.351 - 5.69; p<0.001) more likely to be ZD than those children whose mothers received 4+ antenatal care visits (as per the WHO recommendations). The odds that a child whose mother had less than 4 antenatal care visits was 1.6 times (CI: 1.23 - 2.047; p<0.001) more likely to be ZD than those whose mother had 4+ visits. Regarding the place of delivery, the odds of a child, born at home was 1.3 times more likely to be ZD than those born in an institutional setting, although this association was not statistically significant (p-value = 0.10).

For the postnatal care indicators, the odds of a child who did not receive any check-up within 2 months of being born were 1.2 times more likely (CI: 0.97-1.593; p=0.085) to be ZD than those who did receive the postnatal check-up.

While looking at the predictor variable of households’ engagement with government’s community outreach programs, there was no statistically significant association found between a child’s ZD status and whether its household had met with an Auxiliary Nurse Midwife (ANM) or Lady Health Visitor (LHV) in the past three months. However, the odds of the child being ZD was 1.6 times more likely if the household had not met with an anganwadi worker or Accredited Social Health Activist in the past three months (OR: 1.566; CI: 1.135-2.16; p=006). Similarly, the odds of the child being zero dose was 1.5 times more likely if the mother had not received any benefit from the nearest anganwadi center during pregnancy (OR: 1.529; CI: 1.178-1.986; p=0.001).

## Discussion

For developing effective strategies and initiatives to reach Zero dose (ZD) children, it is important to first identify and characterise ZD households. Through this study, we aimed to develop a profile of a ZD household in the state of Uttar Pradesh, by analysing the household level data from NFHS-5.

As indicated in our analysis, children with ZD immunization status also represent an important marker of general vulnerability, as cumulative consequences of multiple sources of deprivation prevent these children from accessing immunization services. In line with previous studies[14, 15], a statically significant association was observed between the economic status of a household and its status of being ZD, children from the poorer, middle, and richer categories of the wealth index being less likely to being in ZD category, as compared to the children belonging to the poorest wealth quintile. Similarly, children born in Muslim households in the state were more likely to not have received pentavalent dose 1. These findings comply with previous studies conducted in LMICs on correlation between prevalence of ZD children and the demographic indicators such as religion [5] and level of mother’s education [14]. However, in contrast to previous research indicating greater prevalence of ZD children in rural areas in LMICs, [7, 14, 16], our analysis shows that there is an increased risk of being zero dose for children living in urban areas, as urban children were 1.8 times more likely to be in ZD category. This deviation points towards the existence of the ‘urban paradox’[17]; while urban settings generally have better access to health services, urban inequality and exclusion faced by marginalised communities (such as urban slums), can result in many urban residents – including children - miss out and suffer more severe deprivations than their rural peers. Although urban areas generally have higher vaccination coverage, the urban poor are less likely to achieve full immunization compared to the urban average. In some countries, rural residents have better access to healthcare providers due to targeted local interventions [18]. A study conducted in Pakistan also indicated towards higher risk of being ZD for children residing in urban households, compared with those in rural areas [19].

Interestingly, the caste of the child, an important demographic marker in the India socio-cultural context, did not have any significant correlation to the status of receiving pentavalent dose-1, as per our analysis. The present study limits its analysis to NFHS-5 data for the state of Uttar Pradesh. However, similar findings were observed in an analysis to trace aggregate patterns in the numbers of ZD children over time across all states in India, by analysing data from four rounds (1992–2016) of the national health survey. According to the study, the differentials in zero-dose prevalence related to child sex and caste were eliminated over time [5].

Lack of vaccination goes hand in hand with missing out on other health interventions, as unvaccinated children live in households with limited access to routine immunization and other maternal health care services. Studies show that children who received any given vaccine are more likely to also receive other vaccines [16]. Inequalities in accessing immunization services for other pathogen, such as BCG and Polio, was observed to have a statistically significant correlation to the prevalence of ZD children.

Several studies across LMICs [20, 21] and in India [22, 23] have shown that antenatal care (ANC) is a key determinant of childhood vaccination. Like routine vaccination services, ANC requires repeated visits to the health facility and provides an opportunity to guide and educate pregnant women about vaccines for themselves and their newborns. Appropriate and complete ANC including completing 4+ ANC visits, institutional delivery and skilled assistance during childbirth is more likely availed by mothers to whom services are more easily accessible. In compliance, our findings also indicate that households where mothers had not received ANC during pregnancy showed higher prevalence of ZD children. Likewise, place of delivery is an important marker of the ZD prevalence and we found that children born at home are more likely to be in ZD category compared to the children born in an institutional setting. Among children of mothers who did not attend four or more (4+) ANC visits, 20.9% children did not get pentavalent dose 1. For children of mothers who did not attend any ANC visit during pregnancy, zero dose prevalence was comparatively high (57.9%).

In addition to lack of access to health services, our analysis found that ZD households also lack appropriate and sufficient engagement with the system at the community level and limited interaction with community health workers. Households with interactions with frontline health workers like Auxiliary Nurse Midwives (ANMs) and Lady Health Visitors (LHVs) and community health workers like Accredited Social Health Activists (ASHAs) and Anganwadi Workers (AWWs) and who received benefits from and Anganwadi Centre during pregnancy, were more likely to have children who have received pentavalent dose1, as compared to children of mothers who did not have any engagement with health systems. These finding are supported by previous research conducted in India, pointing towards the effectiveness of community health workers and community health interventions in improving uptake of vaccination services [24, 25].

### Strengths and Limitations

The strength of the analysis lies in the robustness of the data source and the meticulous sampling design of the NFHS-5. Using unit-level data from a nationally representative survey provides a comprehensive and reliable basis for drawing conclusions about key determinants of ZD children in the state of Uttar Pradesh. The analytical approach, employing a matched-case-control method reduces the variability and increases the power of analysis and thus allowing us for a nuanced examination of determinants. Despite its strengths, the findings should be interpreted with certain caution. Firstly, the cross-sectional nature of the NFHS-5 data restricts the establishment of causal relationships, making it challenging to infer causation from observed associations.

Additionally, the reliance on retrospective self-reported data, particularly for vaccination information, introduces the potential for recall bias, impacting the accuracy of reported vaccination statuses. Furthermore, the inclusion of all eligible women in the selected household for the survey may introduce potential correlation due to shared environment. Nevertheless, the percentage of such households in the analysed sample was less than 10%. While the National Family Health Survey (NFHS) adopts a uniform and comprehensive sample design representative at the national, state/union territory, and district levels, it still exhibits limitations in capturing the demographic characteristics of certain populations such as slum dwellers and mobile population such as nomadic, de-notified tribes etc.

## Conclusion

This study points towards three major recommendations to effectively reach ZD households. Firstly, interventions to reach ZD children provide an opportunity to address ‘Missed Opportunities’ and bring children and caregivers into contact with health systems. As studies conducted across LMICs highlight [26, 27], there is a need to develop integrated strategies for maternal and child health and immunization services, to efficiently reduce inequalities in health service coverage. Secondly, efforts should be made to strengthen the quality and frequency of engagement with health systems through community health workers for improved knowledge and awareness about routine immunization resulting in improved uptake of vaccination services. Lastly, as the prevalence of ZD household is higher in urban areas in the state of Uttar Pradesh, the focus should be on urban health intervention, to reduce immunization inequalities in the state.

## Declarations

### Consent for publication

Not applicable

## Data Availability

Data used in this study for analysis is publicly available and can be obtained from http://rchiips.org/NFHS/index.shtml and https://nhm.gov.in/index1.php?lang=1&level=2&sublinkid=1332&lid=713

## Acknowledgements

The authors gratefully acknowledge the invaluable contributions of all individuals who supported and facilitated this study.

